# Challenges and opportunities of gap score methods for studying psychopathology resilience and vulnerability

**DOI:** 10.64898/2026.06.13.26355592

**Authors:** Hailey Modi, David AA Baranger, Jared Balbona, Samuel Naranjo Rincón, Aaron J Gorelik, Ryan Bogdan, Janine D Bijsterbosch

## Abstract

**Background:** The widespread prevalence of psychopathology, which affects approximately 50% of the global population, often manifests during adolescence. Understanding why some individuals remain resilient while others experience mental health challenges despite similar environmental risks is essential for developing early interventions. However, past efforts have faced challenges with the retrospective definition of resilience. Here, we aim to address these challenges by quantifying resilience to psychopathology at the individual level.

**Methods:** In the Adolescent Brain and Cognitive Development (ABCD) Study® (N = 11,868), we utilized gradient-boosted tree regression to predict 2-year follow-up psychopathology from 208 Social Determinants of Health features. We used the “gap score” method—the difference between model-predicted and reported psychopathology—to quantify individual differences in psychopathology resilience and susceptibility, defined as the Resilience-Susceptibility Gap (RS-Gap). We validated the RS-Gap against independent 3-year follow-up clinical and quality-of-life outcomes.

**Results:** Collinearity between gap scores and reported symptoms was high (r=-0.84), requiring further correction. Four bias-correction techniques were implemented and compared. After appropriate bias-correction, greater RS-Gap scores were associated with a higher likelihood of poor academic and social outcomes one year later, suggesting that early adaptation to adversity may carry a latent long-term cost.

Conclusions

Dependency between RS-Gap and psychopathology scores is a statistical challenge for gap score resilience methods. Our comparisons demonstrate that correction is mandatory to separate resilience signal from shared variance with psychopathology scores. Findings converged across different bias correction methods, providing a validated framework for using gap scores to identify high-risk developmental trajectories in youth.

## Introduction

The widespread prevalence of psychopathology (i.e., ∼50% of the global population across the lifespan) and its devastating impact on individuals, families, and society (e.g., annual costs of $5 trillion USD in 2019) constitutes a global health problem^1^. Mental health problems typically emerge during adolescence, with ∼25% of their resulting global impact on disability occurring before individuals reach 25 years of age^2^. As such, it is important to understand how individual differences in psychopathology risk emerge at this time due to adolescence being a critical period for development of cortical networks and higher-order processing, as well as subsequent impact on education and future life outcomes^3,4^.

Social determinants of health (SDoH) reflect potentially modifiable environmental and social conditions wherein people are born, live, and age (e.g., socioeconomic status, adversity, neighborhood, pollution, diet). SDoH are among the strongest correlates of psychopathology, accounting for roughly 45% of risk^5^. However, mental health problems following exposure to adverse SDoH are not inevitable; there are vast individual differences in SDoH-related psychopathology risk^6^. Prior work has shown evidence for the psychological construct of resilience, which is hypothesized to contribute to this heterogeneity in outcomes^7–9^. Resilience is largely defined as the successful adaptation to challenging life experiences, resulting in reduced risk of onset and/or less severe psychopathology outcomes as compared to a cohort of peers^10^.

Importantly, an inherent barrier to measuring resilience is that it is a latent construct, defined by phenotypes that are *not* present. Therefore, the majority of quantitative models adopt retrospective investigation in the context of response to a single traumatic event (i.e., PTSD)^11^ or specific exposures (e.g., traumatic event exposure)^12,13^. Although these studies have identified key predictors such as social conflict and genetic liability^10,14^, there is an opportunity to apply this knowledge to prospectively identify risk and resilience at the individual level. Here, we aimed to model the comprehensive effects of SDoH on the development of psychopathology to derive an individualized resilience score.

One promising methodology to quantify resilience and risk on an individual level is to leverage residuals from a machine learning model (so called gap or difference score analysis). The most well-known gap score is that of the Brain Age Gap, which estimates one’s age from neuroimaging-derived features and compares this to actual chronological age to identify relatively premature or delayed brain aging^15–17^. When applied to psychopathology, models are typically designed to predict symptoms instead of age. Prior applications examining psychopathology in childhood cohorts using gap scores have found associations between social response to stress and structural and functional brain changes, as well as effects on future cognition^18,19^.

Although powerful, gap-score approaches have important caveats that need to be addressed to ensure interpretability. The Brain Age literature reports a tradeoff between accuracy and reliability in the calculation of these gap scores^20^. Due to the nature of predictive modeling, many Brain Age models exhibit “regression dilution,” or the tendency of all predictions to skew towards the dataset mean^21^. This systematic bias in prediction error subsequently causes a dependency between the model residuals (i.e., the gap score of interest) and the original prediction target (i.e., age in Brain Age models and psychopathology in resilience models)^22^. Adjustment methods to mitigate this resulting high correlation between the residual and the target variable include regressing the target variable out of the residual, as well as more sophisticated determinations of a corrected prediction^21,23^. We build on prior work by developing and rigorously validating the gap score approach to prospectively quantify individual differences in risk and resilience to psychopathology by testing multiple bias-correction approaches to explore the longitudinal efficacy of our gap score.

We leveraged the Adolescent Brain and Cognitive Development^SM^ Study (ABCD Study®; N = 11,868), an ongoing multisite study of adolescent health and development in the United States^24^, to estimate an SDoH-based Resilience/Susceptibility Gap (RS-Gap) and characterize its correlates. *First*, we entered SDoH features from the 2-year follow-up timepoint into a gradient-boosted tree regression model to predict mental health problems cross-sectionally and explored which SDoH features were most important. *Second,* we calculated RS-Gap scores for individuals by subtracting reported psychopathology from model-predicted psychopathology (see **Fig. 1**). Positive RS-Gap scores reflect relative resilience (i.e., less reported psychopathology than predicted from SDoH features) while negative scores reflect relative susceptibility (i.e., greater reported psychopathology than predicted from SDoH features). *Third*, we validated RS-Gap by testing associations with mental health and quality of life outcomes 1 year later to investigate its utility as a predictive measure. *Fourth,* we implemented multiple statistical methods to determine the robustness of RS-Gap and its ability to capture variation separate from established behavioral assessments.

**Figure 1:**
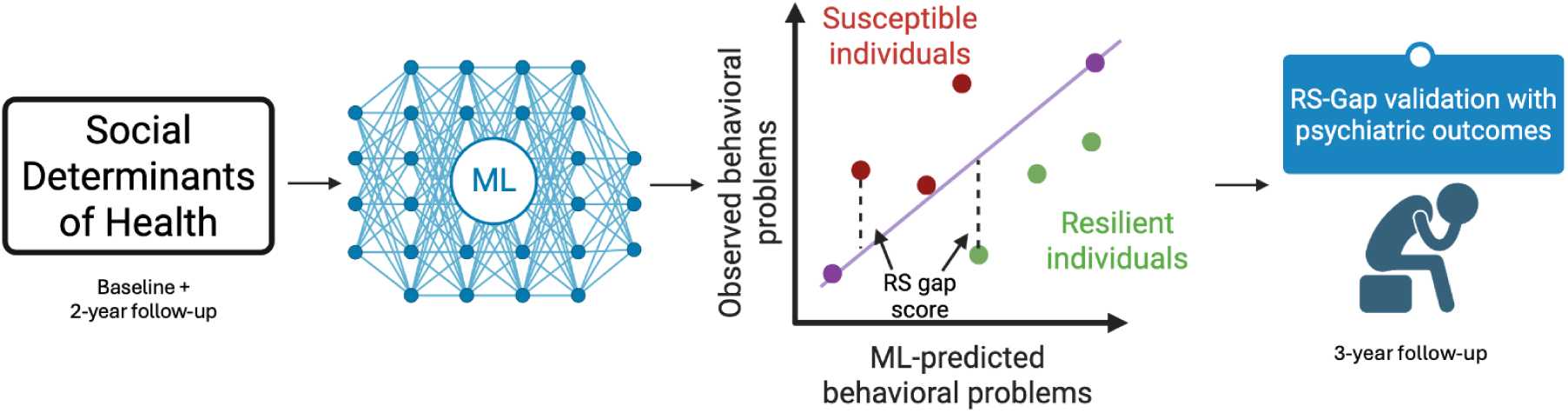
Method Workflow. The RS-Gap score, calculated by taking the difference between machine learning (ML)-predicted and observed psychopathology, will be used to characterize resilience/susceptibility (RS-Gap) in order to test associations with follow-up outcomes at later timepoints. Image created in BioRender.

## Methods

### Participants

The Adolescent Brain and Cognitive Development (ABCD)^SM^ Study is a longitudinal study of complex behavior and biology from middle childhood to late adolescence/young adulthood^25^. The ABCD Study^®^ recruited 11,868 children (ages 8.9 - 11; born between 2005-2009) at baseline (June 1, 2016 - October 15, 2018) from 21 research sites across the United States (https://abcdstudy.org/sites/abcd-sites.html) and includes a family-based design in which twins (n = 2,108), triplets (n = 30), non-twin siblings (n = 1,589), and singletons (n = 8,148) were recruited. All parents or caregivers (10,131 of 11,875 biological mothers [85.3%]) provided written informed consent, and children provided verbal assent to a research protocol approved by a centralized, and each site’s, institutional review board.

Data (release 5.1; https://abcdstudy.org/) from the baseline, 1-year follow-up (1YFU; phenotypic data; n = 11,199; Dates: 8/30/17-3/1/20), 2-year follow-up (2YFU; phenotypic & neuroimaging data; n = 10,066; Dates: 7/30/18-9/27/21), and 3-year follow-up (3YFU; phenotypic data; n = 9,508; Dates: 8/7/19-1/15/22) sessions^1^ were obtained from the National Institute of Mental Health Data Archive (NDA; https://nda.nih.gov/abcd; new data releases will be available on the NIH Brain Development Cohorts Data Hub: https://www.nbdc-datahub.org/). Variable missingness and participant attrition resulted in an analytic sample of 10,266 participants (**Table 1**) for machine learning prediction and 9,327 participants for 3-year follow-up association analyses.

**Table 1:** ABCD Study Sample Characteristics^a^.

| Variable | Total (N = 10266) |
| --- | --- |
| Child variables |  |
| Age, mean (SD), y | 12 (0.7) |
| Girls | 4856 (47.3) |
| Race/ethnicity<sup>b</sup> (n = 10245) |  |
| White | 7798 (76.1) |
| Black/African American | 2019 (19.7) |
| American Indian, Native American, Alaska Native | 356 (3.5) |
| Pacific Islander: Native Hawaiian, Guamanian, Somoan, Other Pacific Islander | 66 (0.6) |
| Asian: Asian Indian, Chinese, Filipino, Japanese, Korean, Vietnamese, Other Asian | 655 (6.4) |
| Hispanic (n = 10737) | 2007 (19.6) |
| Other Race | 655 (6.4) |
| Household income (n = 10101), \$ | |
| <= 49 999 | 3028 (30) |
| 50 000 – 74 999 | 1390 (13.8) |
| 75 000 – 99 999 | 1482 (14.6) |
| 100 000 – 199 999 | 3059 (30.3) |
| >= 200 000 | 1138 (11.3) |

Adolescent Brain and Cognitive Development demographic data at 2-year follow-up.
<sup>a</sup>The estimates reported above refer to the non-winsorized versions of each variable (refer to Supp. 1 for descriptive information of the winsorized data). Unless otherwise specified, the format is number of individuals (percentage).
<sup>b</sup>Race/ethnicity variables were coded as non-mutually exclusive dichotomous variables as reported within ABCD; as such, these numbers do not sum to 100% as participants could be included in multiple categories.

### Resilience/Susceptibility Gap (RS-Gap). SDoH Predicted Psychopathology

To develop a resilience/susceptibility gap (RS-Gap), we first predicted 2-year follow-up (2YFU) child psychopathology based upon Social Determinants of Health (SDoH) features using gradient-boosted (XGB) and random forest regression tree (RF) models that were compared based on model goodness-of-fit, true/predicted score correlations, and across-site variability to identify which model performed better.

***SDoH Input Features***. SDoH variables from baseline and 2-year follow-up (2YFU) sessions were extracted as input features for predictive modeling (F=208; **Table 2**). Data from both baseline and 2YFU sessions were included to leverage the significantly expanded environmental batteries introduced during the follow-up assessments. While features were entered into the machine learning models as a single, unweighted pool without structural partitioning, they are conceptually organized into ABCD-assigned subdomains to facilitate interpretation. These categories include: 1) Family and Socioeconomic Environment (e.g., family environment, parenting metrics, household composition, and parental income/education; n=72), 2) Development and Personal History (e.g., developmental history; n=44), 3) Stress and Adverse Experiences (e.g., severe life events; n=25), 4) Community and Built Environment (e.g., neighborhood cohesion, walkability indices, and health burden metrics; n=24), 5) Peer and Social Dynamics (e.g., peer relationships; n=19), 6) School Context (n=12), 7) Substance Use Environment (n=9), and 8) Air Pollution (n=3). A more granular listing can be found in **Table S1**.

**Table 2:** Social Determinants of Health Input Features.

|  |
| --- |
| **Community** |
| Air pollution (n = 3; satellite-based measures) |
| Population density (NASA Socioeconomic Data and Applications Center) |
| National Walkability Index (EPA) |
| Community cohesion (n = 5; PhenX Toolkit) |
| Neighborhood Safety & Crime (n = 4; PhenX Toolkit) |
| Discrimination Scale (n = 11) |
| Community drug use (n = 2; PLACES) |
| Area Deprivation Index |
| **Development** |
| Biological mother (ABCD Developmental History Questionnaire) |
| Parental age at birth (n = 2; ABCD Developmental History Questionnaire) |
| Twin status (ABCD Developmental History Questionnaire) |
| Vitamin usage and medical visits (n = 2; ABCD Developmental History Questionnaire) |
| Substance exposures (n = 17; ABCD Developmental History Questionnaire) |
| Pregnancy and birth complications (n = 22; ABCD Developmental History Questionnaire) |
| **Family** |
| Family Environment Scale (n = 63; PhenX) |
| **Parenting** |
| Parental Monitoring (n = 5) |
| Household income (ABCD Demographic Survey) |
| Parent/caregiver education (ABCD Demographic Survey) |
| Parent/caregiver marital status (ABCD Demographic Survey) |
| **Peers** |
| Friend and peer prosocial and rule breaking behaviors (n = 4; Peer Behavior Profile) |
| Victimization (n = 9; Peer Experience Questionnaire) |
| Support against substance involvement from close friends (n = 3; Peer Network Health) |
| Number of friends (n = 2; ABCD Youth Resilience Scale) |
| **School** |
| School Risk & Protective Factors (n = 12; PhenX) |
| **Stress** |
| Stressful life events (n = 25; PhenX) |
| **Substance Use Environment** |
| Parental Rules on Substance Use (n = 3) |
| Substance availability (n = 6; Community Risk and Protective Factors) |

**Machine learning model input features comprised social determinants of health metrics at baseline and the 2-year follow-up.** Ns represent the number of individual items from each questionnaire/survey, which are each included as a separate feature. Categories are shown for ease of understanding and are not included in the predictive model.

***Model Prediction: Child Psychopathology.*** SDoH input features were used to predict the 2YFU Child Behavior Checklist (CBCL)^26^ total problem score, which reflects a general index of broad spectrum psychopathology, inclusive of externalizing, internalizing, somatic, and neurodevelopmental problems.

***Machine learning.*** We used regression tree models due to their performance with large-scale, nonparametric, and highly heterogeneous data such as ABCD^27^. We tested the efficacy of both a gradient-boosted tree regression model and a random forest model using the Python packages XGBoost and scikit-learn, respectively^28,29^. XGBoost’s histogram-based gradient boosting regression tree was used due to its ability to handle missing data by using all available data per individual to generate predictions, as well as the non-parametric nature of regression trees lending itself well to large heterogeneous data. The random forest model required listwise deletion of subjects and features with excessive missingness; however, the independent nature of tree building within this method protects more against overfitting. Leave-One-Site-Out Cross-Validation (LOSOCV) was used to account for the multi-site structure of ABCD^30,31^ as well as familial dependencies due to related individuals belonging to the same sites. Nested 5-fold cross-validation within a grid search pipeline was used to generate the best possible tree depth and number of estimators for the regression algorithms (assessed by measuring explained variance, R^2^ value, and mean absolute/squared error). Models generated a predicted 2YFU CBCL score on the held-out testing site (range of n_site_ = 278-934 per cross-validation fold) based on SDoH input features. Hyperparameter settings for the models as well as feature list for the RF model can be found in **Tables S2-3**.

***Feature Importance.*** SHapley Additive exPlanations (SHAP) analysis from the SHAP python package (v0.51.0) was used to characterize feature importance within the best performing model. SHAP analysis estimates the average marginal contribution of each feature (including interactions between features) in the model from each individual prediction instance using the TreeExplainer function optimized for nonlinear decision trees^32^. SHAP values were compared across sites to determine consistency of features used in prediction across each Leave-One-Site-Out Cross-Validation fold.

***Resilience-Susceptibility Gap (RS-Gap) calculation.*** Reported CBCL total problems scores were subtracted from model predicted scores to estimate a continuous RS-Gap for each individual (**Fig. 1**). As such, positive RS-Gap scores reflect relative resilience (i.e., observed behavioral problems lower than predicted by SDoH); negative RS-Gap scores indicate relative susceptibility (i.e., higher behavioral problems than predicted by SDoH).

### RS-Gap validation using independent longitudinal outcome measures

RS-Gap scores were based on SDoH input features measured when children were 9-11 (baseline) and 11-12 (2YFU) and psychopathology outcomes were assessed a year later when children were 12-13 (3YFU). To assess the validity of RS-Gap scores, we estimated RS-Gap associations with psychopathology and related quality of life data (e.g., school performance, psychiatric symptoms).

***Kiddie Schedule for Affective Disorders and Schizophrenia (KSADS) Diagnoses and Symptoms.*** The Kiddie Schedule for Affective Disorders and Schizophrenia assessment (KSADS) is a semi-structured psychiatric diagnostic interview designed for school aged children and adolescents^6^. Disorder diagnoses (e.g. Conduct Disorder, ADHD) as well as individual symptoms associated with each disorder that had prevalence rates of 1% or greater were used in analyses (**Table S4**). KSADS diagnostic data and symptoms were used as the outcome of interest, as opposed to 3YFU CBCL scores, to avoid circularity.

***KSADS Quality of Life Measures.*** We estimated associations between RS-Gap and 7 quality of life factors assessed during 3YFU in the KSADS that had less than 10% missingness in the cohort: 1) detentions/suspensions, 2) drop in grades, 3) repeating a grade, 4) use of mental health or substance abuse services, 5) bullying, 6) having a regular friend group, and 7) having a best friend (**Table S4**).

***Association Analyses.*** Values on continuous outcome variables were standardized and winsorized to ±3 SD prior to analyses to minimize the influence of extreme values^33^. Participants with missing data for 3YFU KSADS measures (n=181) were listwise deleted. All analyses were conducted using linear mixed-effects models (lme4 R [version 4.2.1] package^34^) with random intercepts for site and family to account for their nested structure and dependency. Fixed effects covariates included child sex and age in months at the KSADS to account for sex and age differences in risk^35–37^. Consistent with recently published guidelines on the use of race in biomedical research by the National Academies of Sciences, Engineering, and Medicine^38^, race/ethnicity was not included as a covariate or moderator in analyses.

First, association analyses were used to evaluate the full complement of 27 individual KSADS item outcomes (comprising 20 distinct diagnoses/symptoms + 7 quality of life outcomes). Item-level evaluations were restricted to variables meeting an a priori endorsement threshold of >1% within the sample. False discovery rate (FDR) correction was used to adjust for 27 comparisons. We also clustered individual symptom items into broader binary domains representing any symptom endorsement within that category. This was done to ensure robust, stable estimates. All diagnoses and alcohol use & psychosis symptoms fell below this 1% threshold due to low endorsement rates and were excluded from this step, yielding five final aggregate domains: any ADHD, any Conduct Disorder, any eating disorder, any substance use, and any suicidality symptoms. For each of these domains, participants who endorsed any one or more of the KSADS items included (see **Table S5**) were scored as 1 and participants who did not endorse any of the domain items were scored as 0. Aggregating these items allowed for further validation of the initial item-level phenotypic associations by protecting against sparse data bias and extreme zero-inflation inherent to psychopathology in the ABCD study.

### Bias assessment for gap models

Frequently observed collinearity between gap scores (e.g., our RS-Gap) and the data from which they are derived (i.e., reported CBCL total problems)^20,39,40^ can result in associations that may be attributable to the measure itself rather than the gap score. As a residual, the gap measure is often highly correlated with the target score being predicted, and the degree of this correlation depends on the strength of the relationship between the two variables (i.e., model accuracy). As a result of this collinearity, any association between our resilience metric and an external variable may be driven by the correlated behavioral problems rather than being uniquely related to our operationalized measure of resilience^41^. To address collinearity between RS-Gap and psychopathology, we evaluated four different approaches: 1) model prediction target covariate inclusion, 2) corrected gap scores, 3) removal of highly collinear participants, and 4) extreme group analyses, all described below.

<u>Covariate inclusion.</u> To estimate associations of RS-Gap that are independent of 2YFU CBCL total problems scores (model prediction target), we estimated associations between the RS-Gap and 3YFU KSADS outcomes while including 2YFU CBCL total problems as a covariate. This approach is commonly used in difference score analyses, including gap scores, such as Brain Age^21,41^.

<u>Corrected RS-Gap.</u> As described by De Lange and Cole^42^ as a correction approach for Brain Age, we calculated a corrected ML prediction using the slope and intercept of the relationship between reported and predicted CBCL total problems as follows.

First, a linear regression between predicted and reported score was fit using:

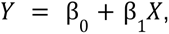

where Y is the model prediction as a function of X, representing the reported CBCL score. This was done to identify if a consistent pattern of error existed between predictions and reported scores. Once β_1_ and β_0_ (the slope and intercept) were calculated, we used these to mathematically adjust prediction to remove patterned bias, based on two previously developed formulae^42^:

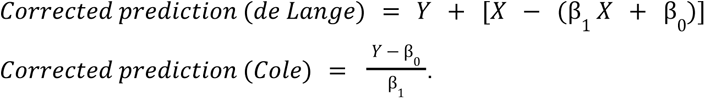

We then calculated corrected RS-Gaps using the corrected predictions, and estimated the correlation of the corrections against reported CBCL score. We also evaluated effects of the corrections on mean absolute error, the correlation between reported and predicted CBCL, and the correlation with the original (uncorrected) RS-Gap score to assess model fit and RS-Gap consistency. These corrected RS-Gap scores were used in the same LME analyses, both with and without 2YFU CBCL. Notably, including CBCL as a covariate should not impact any associations with corrected RS-Gap scores because the colinearity is already addressed. We nevertheless performed associations with and without the CBCL covariate to verify this expected result and mirror all other analyses.

<u>Removal of highly collinear participants.</u> Collinearity between RS-Gap and CBCL was calculated across deciles of the population, and the decile with the largest correlation was removed from subsequent analyses to determine its effect on associations. These LME models were run both with and without 2YFU CBCL as a covariate.

<u>Extreme group analysis.</u> Prior gap analyses have conducted follow-up validation on the ends of the gap distribution because participants with particularly high resilience or vulnerability are likely to provide greater insights than participants with near-zero RS-Gaps^43,44^. Because the decile analysis revealed that it was specifically negative RS-Gaps that were highly correlated with CBCL score, separating this group out allowed us to confirm the effect of collinearity on outcome analyses. Here, we split the participants up into three tertiles based on RS-Gap (N=3,109 each; **Figure S1**). We then separately performed validation analyses in the resilient and susceptible tertiles using the same LME analysis approach, both with and without 2YFU CBCL as a fixed covariate.

## Results

### Model performance

Both the XGBoost (XGB) and random forest (RF) models successfully predicted CBCL Total Problems from SDoH features (XGB_x̂_ R^2^ = 0.25 ± 0.06; RF_x̂_ R^2^ = 0.20 ± 0.07) with similar correlations between reported and predicted CBCL total problems (rs>0.48) as well as RS-Gap distributions (**Figure S2**). Results from the XGB model were used for all subsequent analyses due to: 1) higher performance, 2) less R^2^ variability at each site (**Figure S3**), and 3) the larger sample size (1,600 more participants) and SDoH feature set (48 more SDoH features).

### Feature importance

SHAP analyses revealed that SDoH falling into the family ABCD subdomain were the most influential predictors of CBCL total problems, comprising 40% of the contributions to SHAP score (**Figures 2** & **S4**). This was followed by development (e.g. prenatal residential address, prenatal exposures) and peers/social environment. For the family subdomain, individual importance (absolute SHAP values converted to subject-level contribution to performance) had an average impact of 33.3%, with importance in some individuals rising to 82.1%. Further breakdown of the family subdomain revealed that the top 2.4% of features in this category accounts for approximately 16.2% of the total predictive signal. While there was expected local variation across the 21 sites, the primary prediction drivers (Development and Family subdomains) were globally significant across the entire ABCD Study cohort (**Figure S5**).

**Figure 2:**
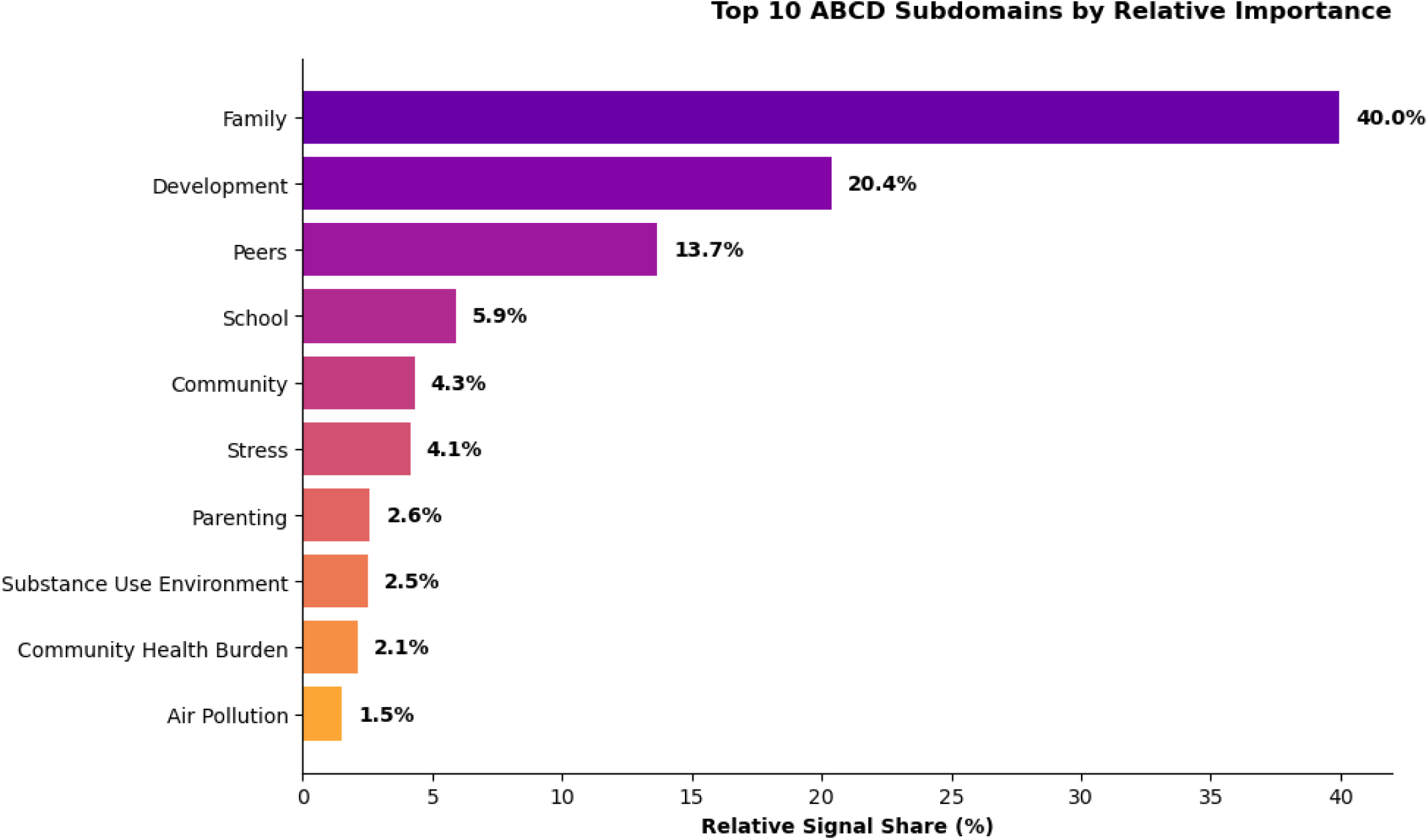
SHAP Analysis. Relative importance of SDoH subdomains. Feature importance is quantified using the mean absolute SHAP (SHapley Additive exPlanations) value across the full analytic cohort. Features are then sorted into subdomains provided as part of the ABCD data release notes. Bars represent the average magnitude of a subdomain’s impact on the model’s output, with percentages indicating the Relative Global Importance—the proportion of the total model signal (sum of all 210 environmental features) attributed to each category of variable. The family domain (i.e. the Family Environment Scale) emerges as the primary driver of the model, accounting for 40% of the total predictive signal for child psychopathology. See **Supplement** for an itemized list of features.

### RS-Gap Descriptives

The RS-Gap distribution was centered on zero with similar numbers of “resilient” (positive values) and “susceptible” (negative values); see **Figure S2A**. 60% of the sample had a positive RS-Gap, meaning predicted CBCL score was greater than observed score (indicating resilience). There was high collinearity between RS-Gap and CBCL Total Problems Score at 2YFU (r=-0.84; **Figure S6**).

### Model validation: Associations with Subsequent Psychopathology and Quality of Life

***Predictive validity and quality of life outcomes.*** The relationship between RS-Gap scores and future outcomes was highly sensitive to whether baseline psychopathology (2YFU CBCL) was included as a covariate. In the uncorrected model, higher resiliency (lower reported psychopathology than predicted) behaved as expected: it was associated with fewer psychiatric symptoms, a lower likelihood of diagnoses, and better school performance. However, introducing the CBCL covariate flipped these results. In this corrected model, positive RS-Gap correlated with worse outcomes across all five psychopathology domains—including increased suicidality, detentions or suspensions, dropping grades, and a lower likelihood of having a close friend group (**Figure 3; Tables S6-7**).

**Figure 3:**
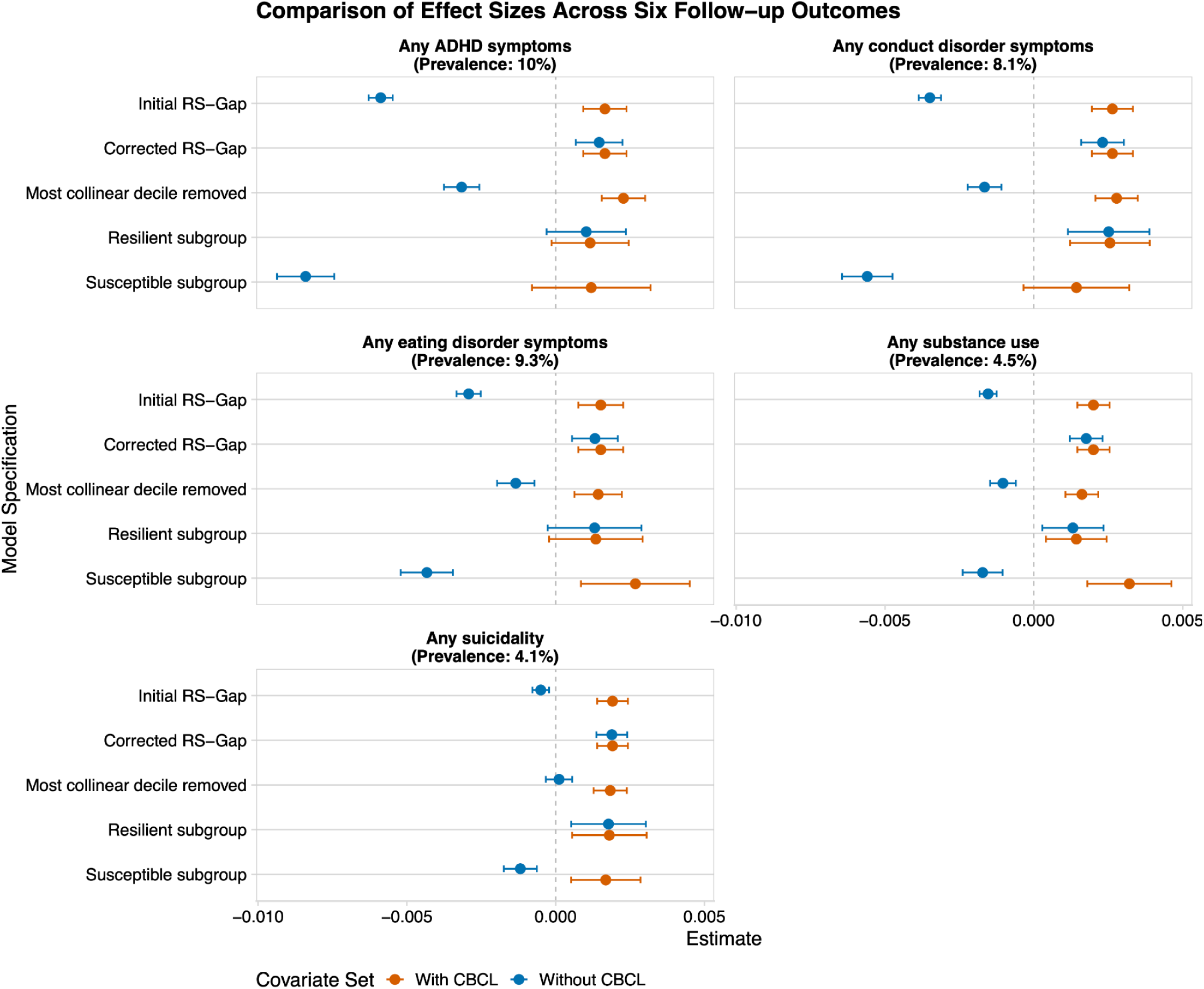
Outcomes Plot. **Comparison of standardized effect size estimates (β) across five model specifications.** Each panel represents a general domain aggregated from the item-level outcomes to represent binary endorsement of any symptom of the following: ADHD, Conduct Disorder, eating disorders, substance use, and suicidality. Points represent the standardized estimate, and error bars represent the 95% confidence intervals. Results are color-coded by covariate specification: Orange points indicate models inclusive of the CBCL covariate, while Blue points indicate models without CBCL. A vertical dashed line at zero denotes the null effect.

***Corrected RS-Gap.*** Next, corrected predictions were calculated using the de Lange & Cole methods, and these were used to generate a corrected RS-Gap for each individual. Distribution of the corrected gaps, as well as correlations and mean absolute error (MAE) as compared to the original prediction and RS-Gaps, can be seen in **Figure S7** and **Table S8**. De Lange’s correction of RS-Gap removed all collinearity with reported CBCL score, while correlation between reported and corrected predicted CBCL increased (r=0.91). Cole’s slope-only correction method also removed collinearity while maintaining the same relationship between reported CBCL score and the corrected prediction (r=0.51). However, while de Lange’s method decreased MAE from 10.3 to 6, Cole’s method caused MAE to increase to 21.8. Prior work has shown that the exclusion of the reported CBCL score in Cole’s method results in a higher variance as compared to the original data^42^, in line with the observed increased MAE. The corrected RS-Gap scores were moderately correlated with the original, uncorrected RS-Gap scores (r=0.54 for both versions), and distributions of the corrected vs. original predictions showed a wider spread of data to compensate for the zero-skewed nature of the CBCL measure (**Figure S7**). While both methods removed the collinearity between RS-Gap and 2YFU CBCL, de Lange’s method resulted in a lower MAE, so subsequent results prioritize de Lange’s method (and matching Cole’s method findings are available in **Table S9**). The de Lange correction resulted in highly similar significance values and effect sizes as the CBCL-as-covariate model, which served to validate the results of both methods **(Figure 3; Tables S10-11)**.

***Removing highest collinearity decile.*** Participants classed as most susceptible (i.e. bottom decile for RS-Gap) were driving the collinearity between the gap score and the target variable (CBCL; **Table S12**). Removing this bottom decile group shifted correlation with CBCL score from r=-0.84 to r=-0.61. When accounting for CBCL, the resulting subsample (N=8,518) showed significant correlation between resilience and drop in grades (β=0.007, p_FDR_=5.2E-28), and lower endorsement of a regular friend group (β=-0.004, p_FDR_=9.43E-12). As for KSADS domains, this cohort indicated positive associations between RS-Gap and all symptomatology domains (β>1.43E-03, p_FDR_<6.32E-04), as seen in **Figure 3**. Item-level results can be seen in **Tables S13-14**.

***Extreme Groups.*** Subsequently, we performed extreme group analyses in which validation was performed separately with the most strongly resilient and the most strongly susceptible groups. Distribution of scores within these subgroups can be found in **Figure S8**. The results of the extreme group analyses broadly confirmed earlier results. Associations between uncorrected RS-Gap controlling for 2YFU CBCL only flipped directionality of associations for the susceptible subgroup, the group with highest collinearity between RS-Gap and CBCL (r=-0.88). The results for the resilient group with low collinearity between RS-Gap and CBCL (r=-0.004) showed the same pattern of poorer quality of life (|β|>0.004, p_FDR_<0.044) and higher likelihood of KSADS symptom endorsement, significant for the domains of conduct disorder, substance use, and suicidality (β>1.31E-03, p_FDR_<0.01), and including CBCL as a covariate did not meaningfully change these results (**Fig. 3; Tables S15-18**). Notably, confidence intervals were large and many associations became nonsignificant compared to results described above that leverage the full (or 90%) sample, reflecting the relatively smaller sample size when performing analyses only in this subset. As such, we observed consistent results of resilience being linked to poorer outcomes when utilizing the original raw RS-Gap scores in the resilient tertile. Importantly, the RS-Gap - CBCL collinearity was not evident in this group such that bias correction (i.e., including CBCL as a covariate) had no impact on the validation results. These findings therefore provide a critically important anchor supporting the validity of the correction methods.

## Discussion

Our study of Social Determinants of Health (SDoH) and resilience/susceptibility to psychopathology revealed two primary findings. First, consistent with a wealth of research linking SDoH factors to psychopathology risk^5^, SDoH variables collectively accounted for a quarter of the total variance in child psychopathology in ABCD. The majority of model performance was attributable to features within the family (40% model performance), development (20% model performance), and peer (14% model performance) domains. In particular, the significant contribution of family dynamics aligns with previous research showing that family cohesion and parental involvement act as strong protective factors against the effects of adverse childhood experiences^45^. Second, we observed that a Resilience-Susceptibility Gap, that reflects a continuum of “susceptibility” (i.e., negative RS-Gap scores indicating more reported psychopathology than predicted by SDoH) and “resilience” (i.e., positive gap scores indicating less reported psychopathology than predicted by SDoH) predicted child psychopathology and quality of life indices 1 year later. Most importantly, our study highlights how collinearity associated with prediction bias can influence the directionality of associations with gap scores and hence their interpretability. As research using measured-prediction gap scores in psychopathology continues to expand^18,46^, it is critical for collinearity between gap scores and measured variables to be carefully considered as is currently being done within other gap score literatures, such as Brain Age^21,23,39,40,42^.

### RS-Gap and Outcomes 1-Year Later: Addressing Prediction Bias Collinearity

Initial models that did not account for reported psychopathology, revealed that “resilience,” as reflected by lower reported psychopathology relative to SDoH-predicted psychopathology (i.e., a positive RS-Gap score), was associated with protection from adverse mental health and quality of life outcomes 1-year later. These findings align with theoretical models (e.g., diathesis stress) of individual differences in stress-related psychopathology risk as well as related psychopathology-gap conceptualizations^18,47^. While intuitively appealing, our analyses addressing the high collinearity between RS-Gap and reported psychopathology, suggest these associations and this interpretation may also be beguiling.

Model prediction scores can be biased toward the dataset mean of the measured variable resulting in collinearity between gap scores and the measured variable being predicted. This occurred in our study as RS-Gap scores were highly correlated (r=-0.84) with reported psychopathology. Analyses considering RS-Gap scores without consideration of reported psychopathology will result in associations that predominantly reflect its shared variance (71%) with measured psychopathology, as well variance (29%) that is unique to RS-Gap. As such, our observed link between RS-Gap and future psychopathology/quality of life 1-year later may be largely driven by variance shared with psychopathology. In other words, as an RS-Gap reflective of resiliency also covaries with less reported psychopathology, the negative association between RS-Gap and psychopathology may simply reflect that prior psychopathology begets future psychopathology.

We used multiple methods (i.e., covariate adjustment, De Lange and Cole correction, removal of the most collinear decile, extreme top/bottom tertile) in an attempt to identify associations of RS-Gap that are not shared with reported psychopathology. All of these adjustments resulted in associations with psychopathology and quality of life metrics 1-year later that were significant in opposing directions to analyses that did not adjust for this collinearity. More specifically, RS-Gap adjusted for reported psychopathology was positively associated with psychopathology and negatively associated with quality of life metrics. As such, scores reflective of relative “resilience,” i.e., less psychopathology reported than predicted by SDoH, were associated with heightened psychopathology and reduced quality of life metrics.

The flip of directionality while adjusting for high collinearity and the intuitive appeal of our uncorrected findings, can reasonably elicit skepticism that our adjusted results may reflect an artifact of collinearity. However, the consistency of findings across models as well as post hoc explanations of these findings increase their plausibility. First, while the majority of variance was shared between RS-Gap and reported psychopathology (71%), a substantial portion (29%) was unique to RS-Gap. Indeed, the variance inflation factor (3.40) was below common thresholds of 5 and 10^48^, suggesting a tolerable level of collinearity that could be included simultaneously within a regression. Moreover, our analyses in the resilient tertile, where collinearity was absent (r=-0.004), revealed a directionality of association that was consistent in both adjusted and unadjusted models suggesting that our adjusted direction of association was recapitulated in a large subsample of the data without collinearity. Finally, we believe this finding is interpretable when considering development in our sample and the duration (i.e., 1 year) of our longitudinal validation analyses. The peak period of psychopathology onset is age 15 with only 10% of psychopathology onsetting prior to age 12^1,3^. As such, the finding that “resilience” as reflected by lower than expected psychopathology at age 10-11 than is predicted by SDoH exposure may reflect censored observations in that these children have not yet made it through developmental periods of psychopathology vulnerability. Hence, in this case, it may be better conceptualized as lower than expected psychopathology as opposed to resilience. It will be important to ascertain whether our findings extend to other developmental periods as the developmental age and duration of follow-up could plausibly induce different directions. For example, among individuals in mid-life who have already been through peak periods of psychopathology risk, lower than expected psychopathology may be associated with protection from psychopathology years later.

### Framework for accounting for collinearity

Meaningfully contending with the relationship between gap scores and target variables is necessary to separate out any actual significant effects that can be attributed to the gap score. Such correction ensures that the created gap measure contains meaningful information beyond what is predicted by the target variable alone. We recommend a robust, two-tiered validation framework:

1. <u>Statistical mitigation.</u> Future designs should include the target variable as a covariate or regress out the linear relationship between gap score and target variable using the de Lange method^42^.
2. <u>Sensitivity anchors via sub-sample stratification</u>. Researchers can combine these statistical corrections with extreme-group analyses to isolate a secondary validation anchor.

Importantly, the degradation of collinearity in the resilient subgroup is not a random occurrence, but a structural consequence of the “floor effect” typical of non-clinical community samples like ABCD. Because severe zero-inflation exists in community psychopathology data, the resilient subgroup is heavily saturated with baseline scores at or near zero. This restriction of variance mathematically attenuates the correlation between the gap score and the target variable. Rather than a limitation, we argue that this variance-restricted sub-sample provides an invaluable, un-collinearized “natural anchor” to verify whether the directionality of the corrected statistical models holds true.

### Strengths, limitations, and future directions

While RS-Gap and similar psychopathology gap scores show promise for identifying high-risk individuals, future work is needed to better understand the effects of collinearity and the extent to which it could affect the interpretation of the data before. Alternative operationalization of resilience using methodologies other than gap score calculation (such as structural equation modeling to identify latent factors) may help determine the reproducibility of these results. While this study leverages longitudinal data to investigate outcomes, the window between the RS-Gap (2YFU) and the validation outcomes (3YFU) is only one year, which may limit our assessment of the long-term correlates of the RS-Gap. The present analyses includes children just entering adolescence, further work using data from older adolescents and teenagers may provide new insights into the longitudinal outcomes for these participants. Moreover, further work is needed to identify the clinical utility of operationalizing resilience using gap analysis^49^, potentially to inform early-intervention strategies.

## Conclusion

Overall, the results of this study provide evidence that leveraging machine learning models to create a gap-score based on social determinants of health (SDoH) can prospectively predict psychopathology outcomes. The resulting “Resilience/Susceptibility Gap” (RS-Gap) showed promise in serving as a robust, individual-level metric of future mental health outcomes. Surprisingly, higher degrees of resilience were linked to poorer behavioral and quality of life outcomes. Our findings additionally highlight that mitigating the unavoidable collinearity between the RS-Gap and psychopathology is a methodological requirement, and we provide a clear framework for future gap-style studies of psychopathology and resilience.

## Data availability

Data used in the preparation of this article were obtained from the Adolescent Brain Cognitive Development (ABCD) Study (https://abcdstudy.org), held in the NIMH Data Archive (NDA). This is a multisite, longitudinal study designed to recruit more than 10,000 children aged 9 to 10 years and follow them over 10 years into early adulthood. The ABCD Study is supported by the National Institutes of Health. A full list of supporters is available at https://abcdstudy.org/federal-partners.html. A listing of participating sites and a complete listing of the study investigators can be found at https://abcdstudy.org/consortium_members/. ABCD consortium investigators designed and implemented the study and/or provided data but did not necessarily participate in analysis or writing of this report. The ABCD data repository grows and changes over time. The ABCD data used in this report came from 10.15154/z563-zd24. The DOIs can be found at http://dx.doi.org/10.15154/z563-zd24.

## Financial Support

This study was funded by F31MH139184 (HM), R01MH128286 (JB), & R01DA054750 (RB). Authors received additional funding support from the National Science Foundation: SNR (2139839), DAAB (R00AA030808).

## Competing interests

The author(s) declare none.

## Supporting information

Tables S2-3, S8, S12, all supplemental figures

## Data Availability

Data used in the preparation of this article were obtained from the Adolescent Brain Cognitive Development (ABCD) Study (https://abcdstudy.org), held in the NIMH Data Archive (NDA).

http://dx.doi.org/10.15154/z563-zd24

## Notes

### Competing Interest Statement

The authors have declared no competing interest.

### Author Declarations

IRB of Washington University in St. Louis gave ethical approval for this work. The IRB protocol number for ABCD is: 201708123.

## References

1. McGrath JJ, Al-Hamzawi A, Alonso J, et al. Age of onset and cumulative risk of mental disorders: a cross-national analysis of population surveys from 29 countries. Lancet Psychiatry. 2023;10(9):668–681. doi:10.1016/S2215-0366(23)00193-1

2. Arias D, Saxena S, Verguet S. Quantifying the global burden of mental disorders and their economic value. eClinicalMedicine. 2022;54:101675. doi:10.1016/j.eclinm.2022.101675

3. Kieling C, Buchweitz C, Caye A, et al. Worldwide Prevalence and Disability From Mental Disorders Across Childhood and Adolescence: Evidence From the Global Burden of Disease Study. JAMA Psychiatry. 2024;81(4):347. doi:10.1001/jamapsychiatry.2023.5051

4. Van Harmelen A. A multilevel systems approach to resilience after childhood adversity. World Psychiatry. 2026;25(2):325–326. doi:10.1002/wps.70070

5. Lean RE, Constantino-Pettit A, Gorham LS, et al. Social Determinants of Health, the developing brain, and risk and resilience for psychopathology. Neuropsychopharmacology. 2026;51(1):185–202. doi:10.1038/s41386-025-02169-1

6. Jirsaraie RJ, Barch DM, Bogdan R, et al. Mapping multimodal risk factors to mental health outcomes. Nat Ment Health. 2025;3(10):1230–1240. doi:10.1038/s44220-025-00500-9

7. Amstadter AB, Myers JM, Kendler KS. Psychiatric resilience: longitudinal twin study. Br J Psychiatry. 2014;205(4):275–280. doi:10.1192/bjp.bp.113.130906

8. Eaton S, Cornwell H, Hamilton-Giachritsis C, Fairchild G. Resilience and young people’s brain structure, function and connectivity: A systematic review. Neurosci Biobehav Rev. 2022;132:936–956. doi:10.1016/j.neubiorev.2021.11.001

9. Kalisch R, Baker DG, Basten U, et al. The resilience framework as a strategy to combat stress-related disorders. Nat Hum Behav. 2017;1(11):784–790. doi:10.1038/s41562-017-0200-8

10. Feder A, Fred-Torres S, Southwick SM, Charney DS. The Biology of Human Resilience: Opportunities for Enhancing Resilience Across the Life Span. Biol Psychiatry. 2019;86(6):443–453. doi:10.1016/j.biopsych.2019.07.012

11. van der Werff SJA, van den Berg SM, Pannekoek JN, Elzinga BM, van der Wee NJA. Neuroimaging resilience to stress: a review. Front Behav Neurosci. 2013;7. doi:10.3389/fnbeh.2013.00039

12. Southwick SM, Charney DS. The Science of Resilience: Implications for the Prevention and Treatment of Depression. Science. 2012;338(6103):79–82. doi:10.1126/science.1222942

13. Kalisch R, Cramer AOJ, Binder H, et al. Deconstructing and Reconstructing Resilience: A Dynamic Network Approach. Perspect Psychol Sci. 2019;14(5):765–777. doi:10.1177/1745691619855637

14. Dahl A, Eilertsen EM, Rodriguez-Cabello SF, et al. Genetic and brain similarity independently predict childhood anthropometrics and neighborhood socioeconomic conditions. Dev Cogn Neurosci. 2024;65:101339. doi:10.1016/j.dcn.2023.101339

15. Cole JH. Multimodality neuroimaging brain-age in UK biobank: relationship to biomedical, lifestyle, and cognitive factors. Neurobiol Aging. 2020;92:34–42. doi:10.1016/j.neurobiolaging.2020.03.014

16. Cole JH, Franke K. Predicting Age Using Neuroimaging: Innovative Brain Ageing Biomarkers. Trends Neurosci. 2017;40(12):681–690. doi:10.1016/j.tins.2017.10.001

17. Franke K, Gaser C. Ten Years of BrainAGE as a Neuroimaging Biomarker of Brain Aging: What Insights Have We Gained? Front Neurol. 2019;10. doi:10.3389/fneur.2019.00789

18. Hettwer MD, Dorfschmidt L, Puhlmann LMC, et al. Longitudinal variation in resilient psychosocial functioning is associated with ongoing cortical myelination and functional reorganization during adolescence. Nat Commun. 2024;15(1):6283. doi:10.1038/s41467-024-50292-2

19. Keller AS, Moore TM, Luo A, et al. A general exposome factor explains individual differences in functional brain network topography and cognition in youth. Dev Cogn Neurosci. 2024;66:101370. doi:10.1016/j.dcn.2024.101370

20. Trafimow D. A defense against the alleged unreliability of difference scores. Cogent Math. 2015;2(1):1064626. doi:10.1080/23311835.2015.1064626

21. Butler ER, Chen A, Ramadan R, et al. Pitfalls in brain age analyses. Hum Brain Mapp. 2021;42(13):4092–4101. doi:10.1002/hbm.25533

22. Smith SM, Vidaurre D, Alfaro-Almagro F, Nichols TE, Miller KL. Estimation of brain age delta from brain imaging. NeuroImage. 2019;200:528–539. doi:10.1016/j.neuroimage.2019.06.017

23. Beheshti I, Nugent S, Potvin O, Duchesne S. Bias-adjustment in neuroimaging-based brain age frameworks: A robust scheme. NeuroImage Clin. 2019;24:102063. doi:10.1016/j.nicl.2019.102063

24. Baskin-Sommers A, Gearing D, Ramduny J, et al. What we have learned about adolescent mental health and where we are going after a decade with the Adolescent Brain Cognitive Development Study. Dev Cogn Neurosci. 2026;78:101686. doi:10.1016/j.dcn.2026.101686

25. Volkow ND, Koob GF, Croyle RT, et al. The conception of the ABCD study: From substance use to a broad NIH collaboration. Dev Cogn Neurosci. 2018;32:4–7. doi:10.1016/j.dcn.2017.10.002

26. Achenbach TM. The Child Behavior Checklist and related instruments. In: Maruish ME, ed. The Use of Psychological Testing for Treatment Planning and Outcomes Assessment. Lawrence Erlbaum Associates Publishers.; 1999:429–466.

27. Klusowski JM, Tian PM. Large Scale Prediction with Decision Trees. J Am Stat Assoc. 2024;119(545):525–537. doi:10.1080/01621459.2022.2126782

28. Chen T, Guestrin C. XGBoost: A Scalable Tree Boosting System. In: Proceedings of the 22nd ACM SIGKDD International Conference on Knowledge Discovery and Data Mining. ACM; 2016:785–794. doi:10.1145/2939672.2939785

29. Buitinck L, Louppe G, Blondel M, et al. API design for machine learning software: experiences from the scikit-learn project. Published online 2013. doi:10.48550/ARXIV.1309.0238

30. Saragosa-Harris NM, Chaku N, MacSweeney N, et al. A practical guide for researchers and reviewers using the ABCD Study and other large longitudinal datasets. Dev Cogn Neurosci. 2022;55:101115. doi:10.1016/j.dcn.2022.101115

31. Garavan H, Bartsch H, Conway K, et al. Recruiting the ABCD sample: Design considerations and procedures. Dev Cogn Neurosci. 2018;32:16–22. doi:10.1016/j.dcn.2018.04.004

32. Lundberg SM, Erion G, Chen H, et al. From local explanations to global understanding with explainable AI for trees. Nat Mach Intell. 2020;2(1):56–67. doi:10.1038/s42256-019-0138-9

33. Kwak SK, Kim JH. Statistical data preparation: management of missing values and outliers. Korean J Anesthesiol. 2017;70(4):407. doi:10.4097/kjae.2017.70.4.407

34. Bates D, Mächler M, Bolker B, Walker S. Fitting Linear Mixed-Effects Models Using **lme4**. J Stat Softw. 2015;67(1). doi:10.18637/jss.v067.i01

35. Modi H, Baranger DAA, Paul SE, et al. Associations between prenatal caffeine exposure and child development: Longitudinal results from the Adolescent Brain Cognitive Development (ABCD) Study. Neurotoxicol Teratol. 2025;107:107404. doi:10.1016/j.ntt.2024.107404

36. Moreau AL, Voss M, Hansen I, et al. Prenatal Selective Serotonin Reuptake Inhibitor Exposure, Depression, and Brain Morphology in Middle Childhood: Results From the ABCD Study. Biol Psychiatry Glob Open Sci. 2023;3(2):243–254. doi:10.1016/j.bpsgos.2022.02.005

37. Paul SE, Hatoum AS, Fine JD, et al. Associations Between Prenatal Cannabis Exposure and Childhood Outcomes: Results From the ABCD Study. JAMA Psychiatry. 2021;78(1):64. doi:10.1001/jamapsychiatry.2020.2902

38. Committee on the Use of Race and Ethnicity in Biomedical Research, Board on Health Sciences Policy, Board on Population Health and Public Health Practice, Board on Health Care Services, Health and Medicine Division, National Academies of Sciences, Engineering, and Medicine. Rethinking Race and Ethnicity in Biomedical Research. (Wilson MR, Beachy SH, Schumm SN, eds.). National Academies Press; 2024:27913. doi:10.17226/27913

39. Smith SM, Vidaurre D, Alfaro-Almagro F, Nichols TE, Miller KL. Estimation of brain age delta from brain imaging. NeuroImage. 2019;200:528–539. doi:10.1016/j.neuroimage.2019.06.017

40. Zhang B, Zhang S, Feng J, Zhang S. Age-level bias correction in brain age prediction. NeuroImage Clin. 2023;37:103319. doi:10.1016/j.nicl.2023.103319

41. Elman JA, Vogel JW, Bocancea DI, et al. Issues and recommendations for the residual approach to quantifying cognitive resilience and reserve. Alzheimers Res Ther. 2022;14(1). doi:10.1186/s13195-022-01049-w

42. De Lange AMG, Cole JH. Commentary: Correction procedures in brain-age prediction. NeuroImage Clin. 2020;26:102229. doi:10.1016/j.nicl.2020.102229

43. Fisher JE, Guha A, Heller W, Miller GA. Extreme-groups designs in studies of dimensional phenomena: Advantages, caveats, and recommendations. J Abnorm Psychol. 2020;129(1):14–20. doi:10.1037/abn0000480

44. Gaser C, Garo-Pascual M, Strange BA. BrainAGE in superagers: cross-sectional and longitudinal analyses in older adults aged 80+ with youthful episodic memory. GeroScience. 2025;47(5):6639–6645. doi:10.1007/s11357-025-01836-x

45. Fritz J, De Graaff AM, Caisley H, Van Harmelen AL, Wilkinson PO. A Systematic Review of Amenable Resilience Factors That Moderate and/or Mediate the Relationship Between Childhood Adversity and Mental Health in Young People. Front Psychiatry. 2018;9:230. doi:10.3389/fpsyt.2018.00230

46. Hammes V, Brosch K, Usemann P, et al. Resilience Beyond Diagnosis: Prospective Neural Correlates of Better-Than-Expected Outcomes in Depression. Psychiatry and Clinical Psychology. Preprint posted online December 30, 2025. doi:10.64898/2025.12.28.25342267

47. Falkenstein T, Sartori L, Malanchini M, Hadfield K, Pluess M. The Relationship Between Environmental Sensitivity and Common Mental-Health Problems in Adolescents and Adults: A Systematic Review and Meta-Analysis. Clin Psychol Sci. 2026;14(2):135–157. doi:10.1177/21677026251348428

48. Kutner MH, Nachtsheim CJ, Neter J, Li W, eds. Applied Linear Statistical Models. 5. ed., internat. ed. McGraw-Hill; 2005.

49. Schulz MA, Siegel NT, Ritter K. Brain-age models with lower age prediction accuracy have higher sensitivity for disease detection. Lewis LD, ed. PLOS Biol. 2025;23(10):e3003451. doi:10.1371/journal.pbio.3003451

