## Supplementary material for "Challenges and opportunities of gap score methods for studying psychopathology resilience and vulnerability": Tables S2-3, S8, S12, all supplemental figures

**Table S2**

**Prenatal Environment**

Vitamin usage and medical visits (n = 2; ABCD Developmental History Questionnaire)^29,30^

Air pollution (n = 3; satellite-based measures)^31,32^

Substance exposures (n = 10; ABCD Developmental History Questionnaire)

**Built Environment**

Neighborhood Safety & Crime (n = 4; PhenX Toolkit)^33,34^

Population density (NASA Socioeconomic Data and Applications Center)

Area Deprivation Index^35^

National Walkability Index (EPA)

Substance availability (n = 6; Community Risk and Protective Factors)^36,37^

Community drug use (n = 2; PLACES)^38^

School Risk & Protective Factors (n = 12; PhenX Toolkit)^39,40^

**Immediate Environment**

Number of friends (n = 2; ABCD Youth Resilience Scale)^41^

Household income (ABCD Demographic Survey)

Parent/caregiver education (ABCD Demographic Survey)

Parent/caregiver marital status (ABCD Demographic Survey)

Parental Monitoring (n = 5)^42,43^

Parental Rules on Substance Use (n = 3)^44,45^

Family Environment Scale (n = 63; PhenX)^46^

Post-Traumatic Stress Disorder (KSADS)^47^

Victimization (n = 9; Peer Experience Questionnaire)^48,49^

Friend and peer prosocial and rule breaking behaviors (n = 1; Peer Behavior Profile)

Support against substance involvement from close friends (n = 3; Peer Network Health)^50^

**Adverse Experiences**

Stressful life events (n = 25; PhenX)^51,52^

Discrimination Scale (n = 3)^53,54^

**Input features for the Random Forest model** (n=171). Feature and participant numbers had to be finetuned for the random forest model because the algorithm does not accept missing data. In order to balance sample size and feature set size, particulate matter pollution, premature birth, and age of biological parents were not included in the random forest regression due to excessive missingness (N_missing_=261-737). This resulted in N = 8,666 participants with complete data on 171 features for the random forest model, encompassing the largest number of participants for which the largest number of features were available.

**Table S3**

| **Random Forest Regression** | | **Gradient-Boosted Tree Regression** | |
| --- | --- | --- | --- |
| Hyperparameter | Range | Hyperparameter | Range |
| Maximum depth | 5, 10, 20, 40, None | Maximum depth | 2, 4, 6 |
| Maximum features | 1, 5, log2, sqrt, None | # of estimators | 50, 100, 200 |

To determine optimized hyperparameters, both machine learning models used a grid search within the estimator pipeline. Selected features and ranges were based on scikit-learn and XGBoost documentation for setting up an ideal grid search. Maximum tree depth refers to the maximum number of nodes from the root node to the leaf node of a given decision tree. Maximum features in a tree structure is the number of features each node will consider in order to create the next split. Because RF and GBM are tree algorithms involving many decision trees, parallelized and sequentialized respectively, the n_estimators parameter can set a limit on the number of individual trees within the model.

**Figure S1**

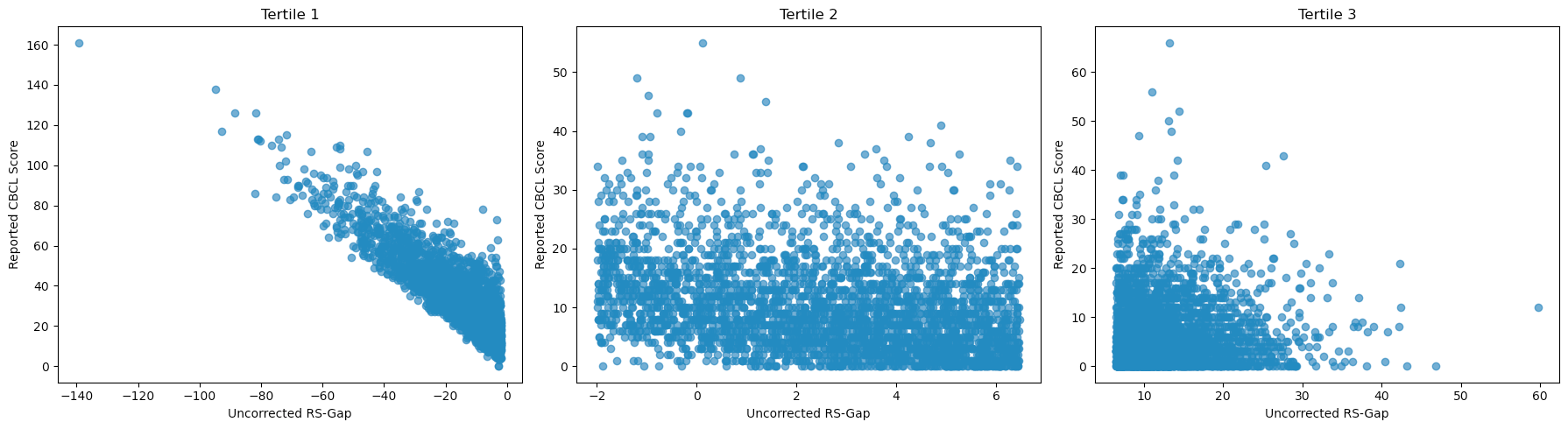

**Stratification of RS-Gap scores into tertiles for collinearity visualization.** Visual comparison of the RS-Gap vs. true 3YFU scores for susceptible (Tertile 1), middle (Tertile 2), and resilient (Tertile 3) groups illustrates the differential correlation patterns across the RS-Gap distribution.

**Figure S2**

A

B

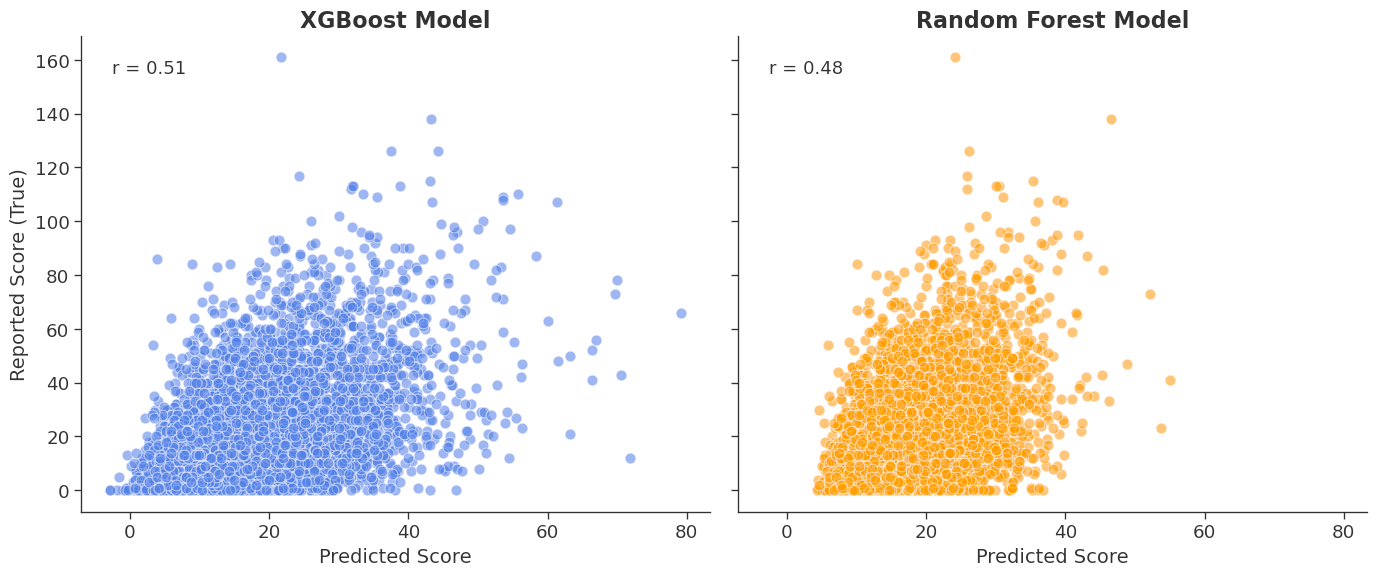

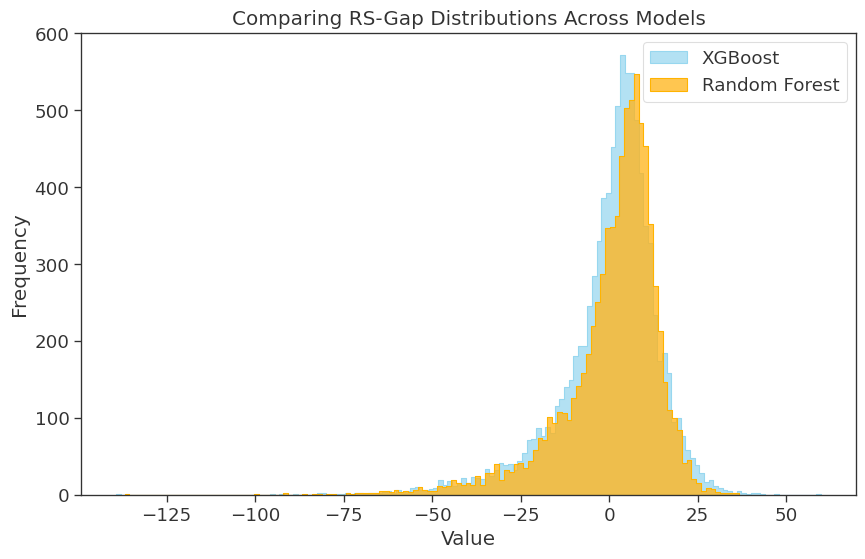

**Panel A: Comparison of Resiliency Susceptibility-Gap (RS-Gap) Distributions.** Overlapping histograms show the distribution of RS-Gap for XGBoost (blue; mean 0.05 ± 14.5; range:-139.2 - 59.8) and Random Forest (orange; mean 0.54 ± 14.3; range: -136.9 - 36.8) models. The "step" outline format highlights the higher frequency of near-zero residuals in the XGBoost model compared to Random Forest. **Panel B: Model Correlation and Prediction Performance.** Side-by-side scatterplots illustrate the relationship between reported CBCL total problems and Social Determinants of Health model-predicted CBCL Total Problem scores. Both models demonstrate moderate positive correlations (r≥0.48).

**Figure S3**
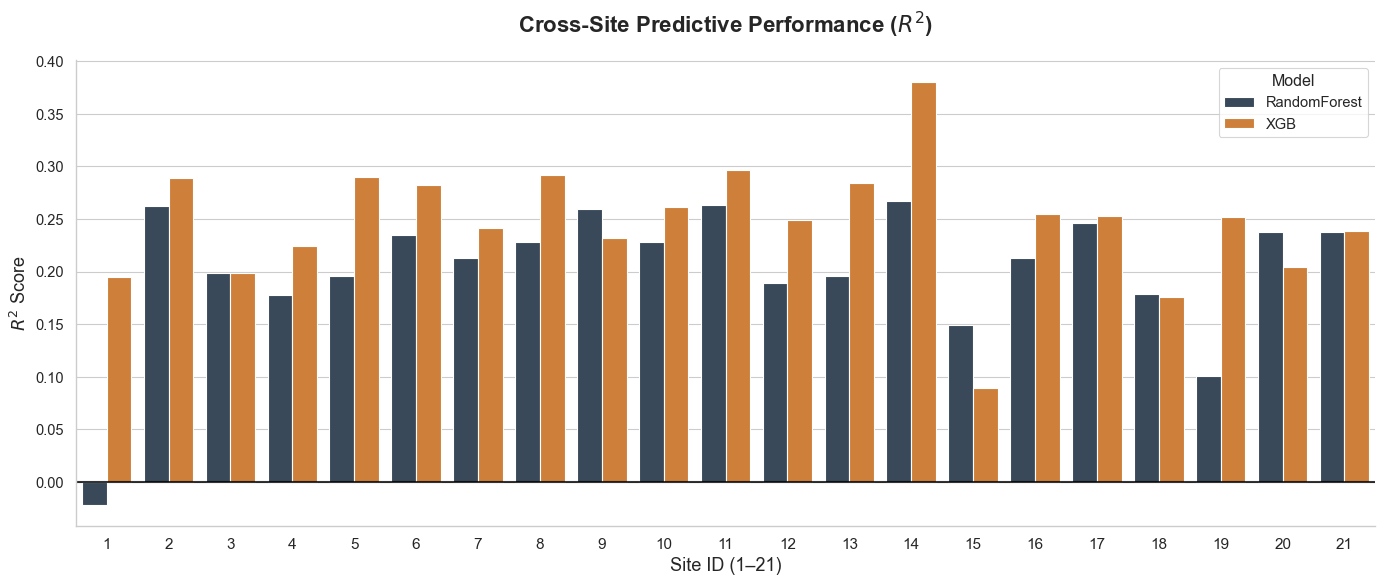

**Predictive Accuracy Across 21 Independent ABCD Sites.** Cross-validated R^2^ scores by study collection site (i.e., 1-21) for XGBoost (orange; R^2^ mean±SD = 0.25±0.06; range 0.08-0.38 and Random Forest (navy; R^2^ mean±SD = 0.20±0.07; range -0.02-0.27) models. The horizontal baseline at y=0 represents the performance of a null model; the single bar below the horizontal baseline indicates where the model failed to outperform a simple mean prediction. However, removing site 1 from the RF prediction model did not meaningfully improve goodness-of-fit metrics.

**Figure S4**

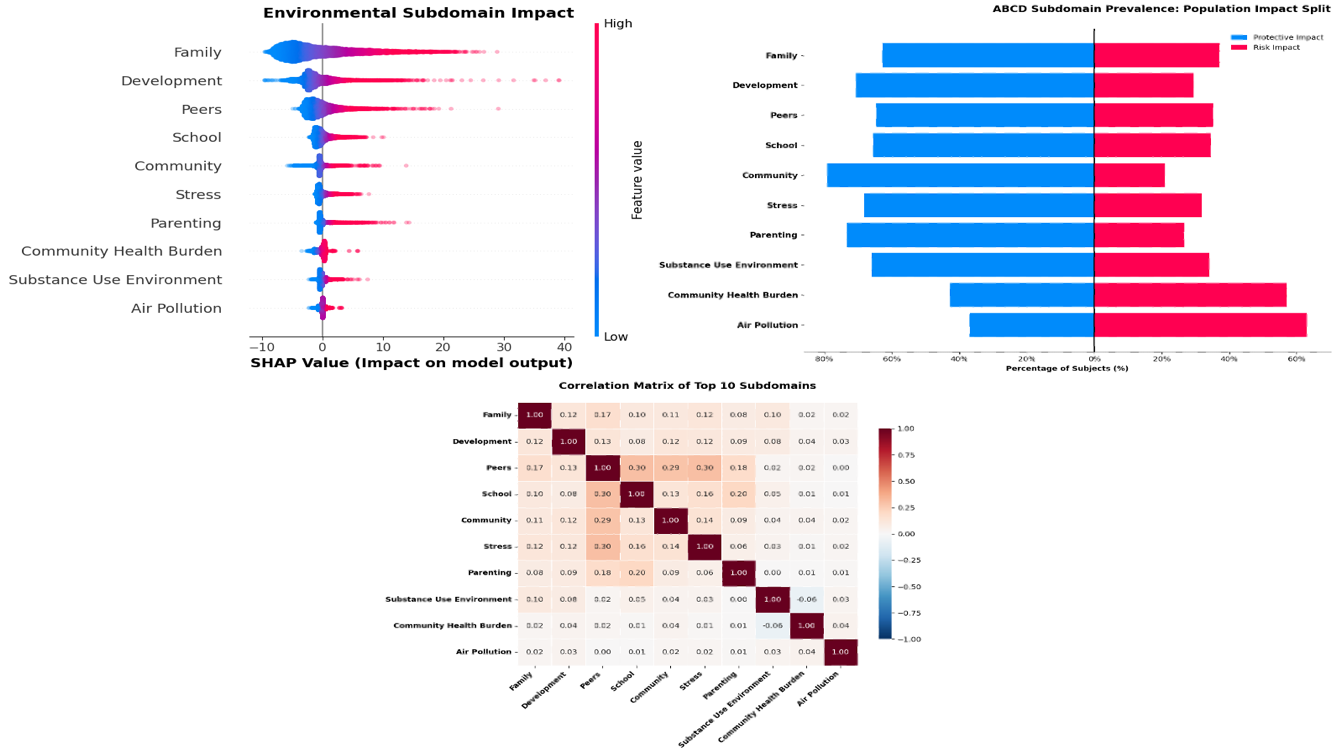

C

A

B

**Characterizing the Predictive Architecture and Geographic Stability of the RS-Gap.** *A:* SHAP summary plot showing the impact of specific feature values on model output. Blue points indicate lower feature values (protective), while red indicates higher values (risk). *B:* Population impact split shows the percentage of subjects for whom each subdomain exerted a protective (blue) versus risk (red) influence. *C:* Correlation matrix showing the independence of the top 10 environmental subdomains, confirming that the predictive signal is not driven by high inter-correlation between environmental categories.

**Figure S5**

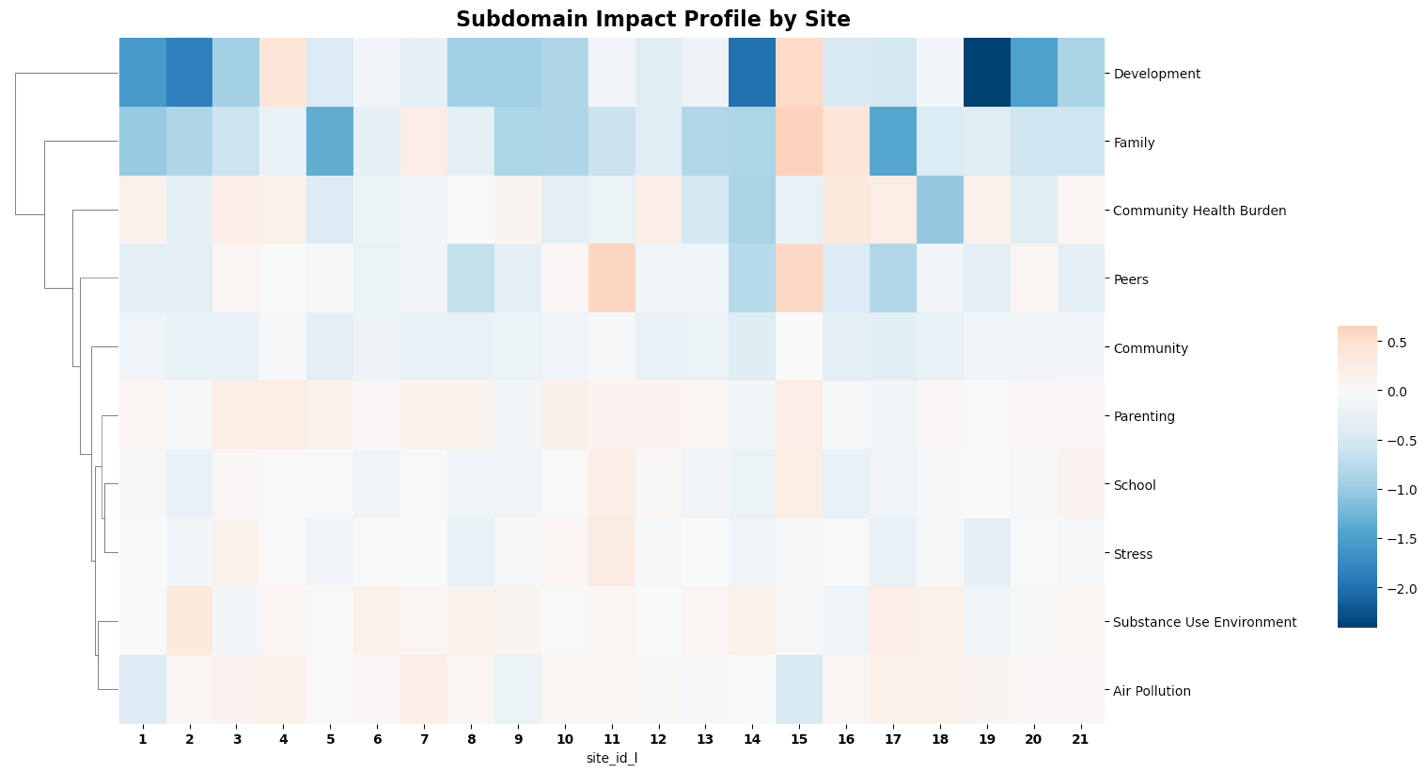
**Site-wide consistency of environmental subdomain impacts across ABCD study locations.** Heatmap illustrating the mean absolute SHAP values (global importance) for the top 10 environmental subdomains, stratified by data collection site (N=21 sites). Minimal site-by-site variability confirms that the environmental determinants of the resilience gap are generalizable across the national cohort rather than being driven by site-specific artifacts or localized population characteristics.

**Figure S6**

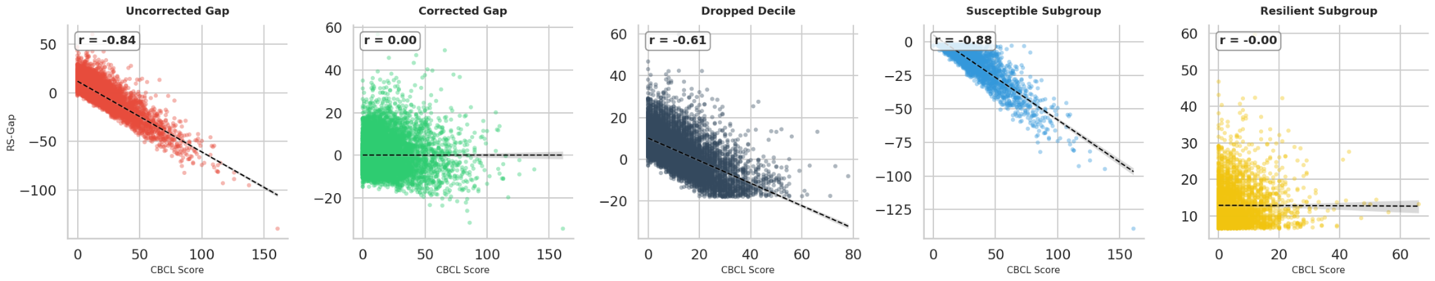

**Evaluation of collinearity across bias assessment methods.** Individual panels display the relationship between clinical symptom severity and resilience gap metrics across various stages of adjustment. From left to right: (1) Uncorrected Gap (r=−0.84), showing severe regression-to-the-mean artifacts; (2) Corrected Gap (r=0.00), showing successful linear decorrelation across the sample; (3) Dropped Decile (r=−0.84), demonstrating the correlation pattern remaining after removing the most skewed decile; (4) Susceptible Subgroup (r=−0.88), representing the isolated bottom tertile; and (5) Resilient Subgroup (r=−0.00), showing a decoupled relationship within the top tertile. Dashed lines represent ordinary least squares linear trendlines with shaded 95% confidence intervals.

**Figure S7**
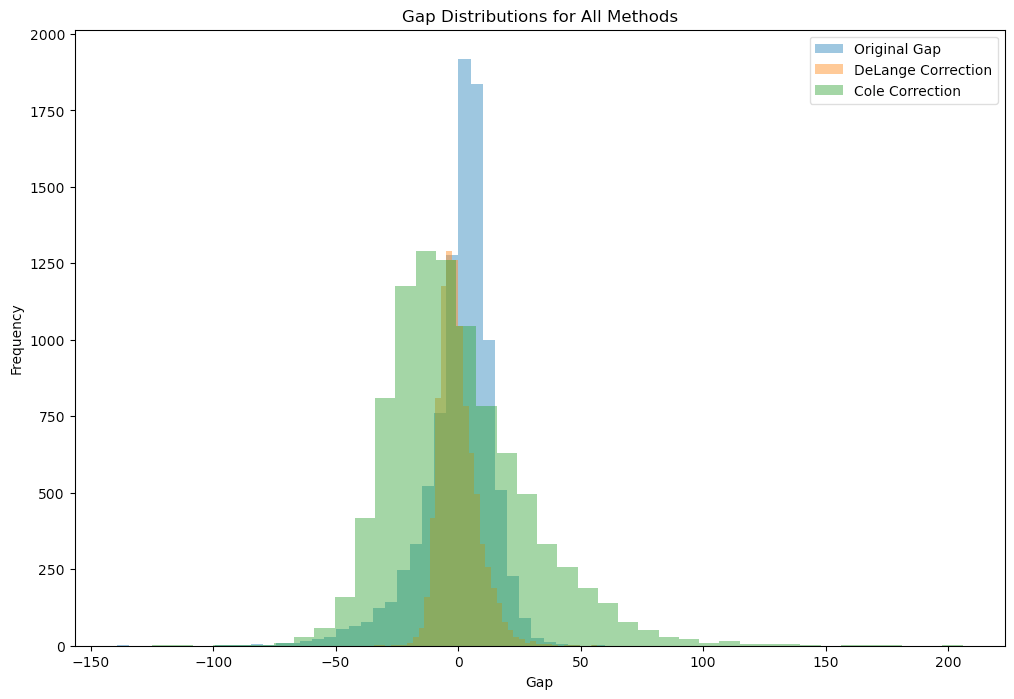

**Comparison of DeLange & Cole Corrections to Original RS-Gap Distribution.** Overlapping histograms show the distributions of the initial RS-Gap as well as the two alternate RS-Gaps calculated from corrected predictions. The DeLange distribution ranged from -34.4 - 56.7 (mean 0 ± 7.84), while the Cole correction resulted in a range of -124.9 - 205.9 (mean 0 ± 28.5). Note that the range of the original RS-Gap is -139.2 - 59.8 (mean 0.05 ± 14.5), as seen in **Figure S2A.**

**Table S8**

|  | **Original RS-Gap** | **DeLange Correction** | **Cole Correction** |
| --- | --- | --- | --- |
| **Predicted to reported CBCL correlation** | 0.51 | 0.91 | 0.51 |
| **RS-Gap to CBCL correlation** | -0.84 | 0 | 0 |
| **Mean absolute error (MAE)** | 10.35 | 6.00 | 21.79 |
| **Original RS-Gap to corrected gap correlation** | 1.0 (by definition) | 0.54 | 0.54 |

**Corrected RS-Gap metrics as compared to initial RS-Gap.** Implementing both statistical corrections resulted in an increase in predicted-reported *r*, as well as negation of the collinearity between RS-Gap and CBCL score. MAE decreased using the DeLange method, while it substantially increased using the Cole method, in line with the higher variance the Cole statistical correction results in. Both corrections were moderately correlated with the original RS-Gap.

**Table S12**

| **Decile [Score Range]** | **Correlation** |
| --- | --- |
| 1 [-139.25, -17.98] | -0.85 |
| 2 [-17.98, -8.35] | -0.35 |
| 3 [-8.34, -3.16] | -0.14 |
| 4 [-3.14, 0.14] | -0.10 |
| 5 [0.15, 2.81] | -0.09 |
| 6 [2.81, 5.00] | -0.07 |
| 7 [5.00, 7.25] | -0.04 |
| 8 [7.25, 10.04] | -0.02 |
| 9 [10.05, 14.44] | 0.07 |
| 10 [14.45, 59.78] | 0.05 |

**Gap Range and Model Correlation Divided Into Deciles.** The study population was partitioned into ten equal-sized deciles (n=809) based on the calculated RS-Gap values to assess the stability of model performance across the data distribution. For each decile, the numerical range (Min–Max) of the gap is provided alongside the Pearson correlation coefficient (r) between the gap and reported scores. Decile 1 is characterized by the most negative gap values, the widest numerical range among all partitions, and the highest correlation coefficient.

**Figure S8**

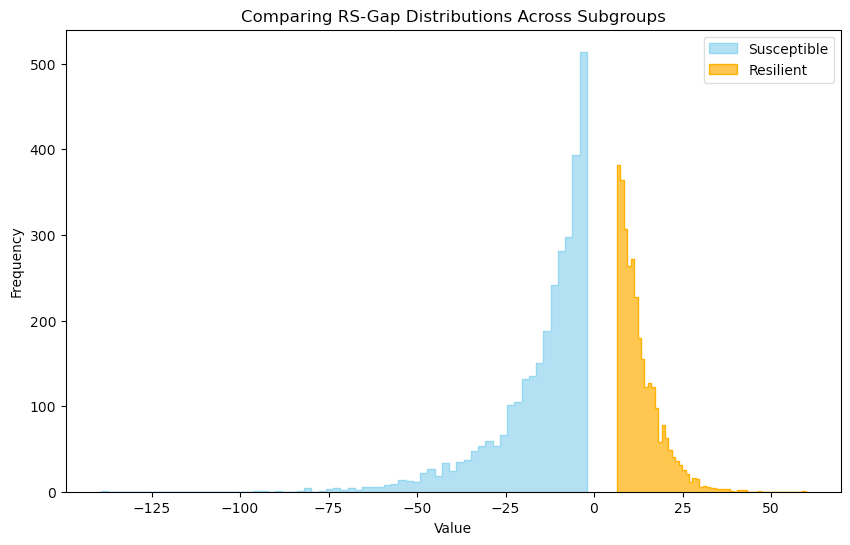

**RS-Gap Score Distributions for Extreme Group Analysis.** Visualization of RS-Gap scores for the most strongly resilient and susceptible participants. The histogram shows the frequency of values within each validation subgroup.
